# Negligible Risk of the COVID-19 Resurgence Caused by Work Resuming in China (outside Hubei): a Statistical Probability Study

**DOI:** 10.1101/2020.03.26.20044271

**Authors:** Kedong Zhao, Cheng Long, Yan Wang, Tieyong Zeng, Xinmiao Fu

**Affiliations:** Provincial University Key Laboratory of Cellular Stress Response and Metabolic Regulation, College of Life Sciences, Fujian Normal University, Fuzhou City, Fujian Province 350117, China; Department of Orthopaedic, Sichuan University West China Hospital, Chengdu City, Sichuan Province 610041, China; Department of Mathematics, The Chinese University of Hong Kong, Shatin, NT, Hong Kong 99999, China

**Keywords:** COVID-19, SARS-CoV-2, coronavirus, COVID-19 resurgence, risk assessment, work resuming

## Abstract

The COVID-19 outbreak in China appears to reach the late stage since late February 2020, and a stepwise restoration of economic operations is implemented. Risk assessment for such economic restoration is of significance. Here we estimated the probability of COVID-19 resurgence caused by work resuming in typical provinces/cities, and found that such probability is very limited (<5% for all the regions except Beijing). Our work may inform provincial governments to make risk level-based, differentiated control measures.

---

The outbreak of 2019 novel coronavirus diseases (COVID-19) has dramatically impacted on China and also starts to hit the world ^1, 2^. Along with a significant decrease of daily new confirmed cases from over 3000 to less than 100 (19 on 9 March 2020) ^3^, China (outside Hubei) has entered a new stage of epidemic prevention and control coupled with a stepwise restoration of social and economic operations. The rapid return to full productivity is critical to China and also to the world that urgently needs the material goods including personal protective equipment against COVID-19 infection ^4^. Rational risk assessment for the COVID-19 resurgence upon such economic restoration is of significance. Here we estimated the probability of COVID-19 resurgence caused by work resuming in typical provinces/cities (refer to **Table 1**) that were most affected by the outbreaks and/or are most economically important in China.

**Table 1.** Probability of COVID-19 resurgence after working resuming and school reopening ^a^

| Province /cities | New cases | No. of cumulative cases | Population (10<sup>4</sup>) | No. of Enterprises (10<sup>4</sup>) | Probability (%) |  |
| --- | --- | --- | --- | --- | --- | --- |
|  |  |  |  |  | mild<sup>b</sup> | strict<sup>b</sup> |
| Guangdong | 2 | 1353 | 11346 | 535.1 | 3.8 | 1.1 |
| Henan | 0 | 1272 | 9605 | 154.2 | 0.0 | 0.0 |
| Zhejiang<sup>b</sup> | 0 | 1215 | 5737 | 224.5 | 0.0 | 0.0 |
| Hunan | 1 | 1018 | 6898 | 83.5 | 0.6 | 0.2 |
| Jiangxi | 0 | 935 | 4647 | 83.8 | 0.0 | 0.0 |
| Anhui | 0 | 990 | 6323 | 141.1 | 0.0 | 0.0 |
| Shandong | 3 | 759 | 10047 | 261.3 | 3.2 | 1.0 |
| Jiangsu<sup>b</sup> | 0 | 631 | 8050 | 347.9 | 0.0 | 0.0 |
| Fujian | 0 | 296 | 3973 | 138.8 | 0.0 | 0.0 |
| Beijing<sup>c</sup> | 3 | 429 | 2153 | 161.6 | 6.8 | 2.0 |
| Shanghai | 1 | 344 | 2423 | 220.8 | 3.7 | 1.1 |
| Guangzhou | 1 | 347 | 1490 | 127.7 | 3.5 | 1.0 |
| Shenzhen<sup>b</sup> | 0 | 419 | 1302 | 201.9 | 0.0 | 0.0 |

<sup>a</sup> Steps for probability calculation are presented in **Table S2**.
<sup>b</sup> There were no newly confirmed COVID-19 cases in these regions from 25 February to 9 March such that the final probability of resurgence is zero. NA: not available
<sup>c</sup> The secondary attack rate was set as 10% and 3% under mild and strict personal protection conditions, respectively, by referring to the estimates on family clusters<sup>4</sup>.

Risk assessment for work resuming is based on several assumptions as follows. First, a period of the past 14 days was set as a reference for calculation, given that the incubation period of COVID-19 ranges from 1-14 days with the mean period of 5-6 days ^4^. Second, potential infection in the coming week is proportional to the number of newly confirmed COVID-19 cases in the past 14 days. Third, only locally generated cases in the past 14 days are counted while imported cases are omitted (Note: all passengers entering China from foreign countries are required to be quarantined for 14 days, and would be subjected to COVID-19 test if necessary ^5^). Forth, the secondary attack rate of COVID-19 in enterprise clusters from an infected but not yet identified case to healthy persons, if not less, is comparable with that in family clusters, which ranges from 3%-10% ^4^. Fifth, there is one cluster of health event in each enterprise every day and the average cluster size is assumed as 10 persons.

Under the above assumptions, we collected the data of new COVID-19 cases in the past 14 days (from 28 February to 12 March; refer to **Table S1**) in each area and also the population size and numbers of enterprises in 2019. Estimation of the probability of COVID resurgence was performed step by step, as detailed in **Table S2**. Results (**Table 1**) indicate that i) under mild and strict protective conditions, probability of COVID-19 resurgence in the coming week (from 13 March to 19 March) ranges from 0.6%-6.8% and from 0.2%-2.4%, respectively; ii) In several areas (e.g., Zhejiang, Jiangsu and Shenzhen) probability is zero due to the absence of new cases in the past 14 days.

In summary, the probability of COVID-19 resurgence upon working resuming is very limited or even negligible. The probability may be updated weekly or daily by referring to the new cases in the past 14 days. Our work may provide guidance for provincial governments to make risk level-based, differentiated control measures, by which economic operations are effectively restored and the potential risk of COVID-19 resurgence is strictly controlled.

## Data Availability

All data are included in the manuscript

## Acknowledgments

This work is support by the National Natural Science Foundation of China (No. 31972918 and 31770830 to XF). We declare no competing interests.

